# Family Planning Performance in 113 Countries Using a New Composite Performance Index

**DOI:** 10.1101/2021.04.11.21255259

**Authors:** Aalok Ranjan Chaurasia

## Abstract

In this paper we measure family planning performance in 113 countries of the world using a composite performance index that takes into account both the met demand for family planning and the composition of the met demand. The composite performance index used in the present paper is an improvement over the existing approaches of measuring family planning performance. The analysis reveals that family planning performance varies widely across countries and it remains either poor or very poor in more than 40 per cent countries. There is no country where the performance is very high. Moreover, there are many countries where the family planning performance appears to have deteriorated over time. There are only a few countries where family planning performance has improved in recent years. The analysis calls for increased investment in family planning to meet the target set under the Sustainable Development Goals.

## Introduction

In this paper, we analyse family planning performance in 113 countries on the basis of the composite family planning performance index proposed by Chaurasia (2020) which conveys summary information about progress in family planning and signal priorities for improving delivery of family planning services. The purpose is to present the ‘big picture’ by offering a rounded assessment of the family planning performance in a simple yet convincing manner that appeals to policy makers, program managers and even to the common people. The family planning performance index used in this paper to assess family planning performance is an improvement over the existing approaches of measuring family planning progress. It takes into account, separately, met demand for modern spacing methods, met demand for permanent methods and method-mix or the composition of the met demand to assess family planning performance.

Family planning performance has commonly been measured in terms of the contraceptive prevalence rate (CPR) because of the strong negative relationship between CPR and total fertility rate (TFR) on the basis of cross-country data (Bongaarts, 1978; Bongaarts and Potter, 1983; Ross and Mauldin, 1996; Jain, 1997; Tsui, 2001; Stover, 1998; United Nations, 2020). Srinivasan (1988) has, however, argued that as one goes down the level of aggregation, variation in CPR explains less and less of the variation in TFR. There are also studies that show inconsistency in the relationship between CPR and TFR, especially in sub-Saharan Africa (United Nations, 2020; Westoff and Bankole, 2001; Adamchak and Mbizvo 1990; Bongaarts 1987; Thomas and Mercer 1995; Jurczynska, Kuang, and Smith 2016; Jain et al, 2014).

Measuring and monitoring family planning performance through CPR, however, has many limitations. First, a proportion of married/in-union women of reproductive age may not be using any family planning method because they either want a child or are sterile and this proportion varies from population to population. A universal upper limit of CPR, therefore, is difficult to establish. Second, CPR does not take into account the distribution of family planning users by different family planning methods or the method-mix. A consideration to method-mix is important as effectiveness of different family planning methods in preventing birth is different and use of different family planning methods is closely linked with family building strategies. This implies that method-mix is essentially different in women at different stages of family building process. It has, therefore, been emphasized that family planning performance measurement should not be limited to just counting the number of women using any family planning method. Rather, family planning performance measurement should also take into the range and types of family planning methods being used (United Nations, 2019).

Recently, the proportion of married/in-union reproductive age women whose demand is satisfied with a modern family planning method or the met demand for family planning has been advocated to measure family planning progress (FP2020, 2018). This indicator is one of the indicators selected to monitor the progress towards Sustainable Development Goal 3 of the United Nations 2030 Sustainable Development Agenda (United Nations, 2015). However, this indicator also has limitations in the context of measuring family planning performance. First, it does not distinguish between the met demand for modern spacing methods and the met demand for permanent methods. This distinction is important as the context of using a modern spacing method is essentially different from that of using a permanent method. A permanent family planning method can be used only when the family building process is complete. If the family building process is not complete, use of permanent methods is out of question. Second, the met demand for family planning is not the simple addition of the met demand for modern spacing methods and the met demand for permanent methods. Third, the met demand for family planning ignores the implicit but very strong assumption about the perfect substitutability between the met demand for modern spacing methods and the met demand for permanent methods. From the perspective of performance measurement, this assumption has little justification. The shortfall in the met demand for modern spacing methods cannot be substituted by the high met demand for permanent methods. Finally, like the CPR, the met demand for family planning is also a crude indicator as it does not take into consideration the method-mix or the proportionate distribution of family planning users by different family planning methods.

Given the limitations of existing approaches, we adopt a two-dimensional approach to measure family planning performance in the context of meeting the diverse family planning needs of women. The first dimension, obviously, is the met demand for family planning. We, however, make a distinction between the met demand for spacing births and the met demand for limiting births as the context of using a modern spacing method is essentially different from the context of using a permanent method. The second dimension that we consider in measuring family planning performance is the method-mix or the proportionate distribution of family planning users by different modern family planning methods. The incorporation of the dimension of the method-mix in measuring family planning performance is important from different perspectives. Method-mix reflects the method choice which is a key principle in both quality of care and rights-based family planning. Method choice is also a guide for optimal delivery of family planning services (WHO, 2014). Method mix also reflects both supply and demand for family planning services. On the supply side, method-mix is optimized when a range of family planning methods are available and accessible to women to meet their diverse family planning needs. Method-mix is also influenced by the demand for different family planning methods including individual and societal preferences and the basic orientation of family planning services delivery system, particularly, organized family planning services. Demand for different family planning methods may also be influenced by side effects that are associated with different methods. Although, interest in method-mix dates back to 1980s (Johnson, 1984; Snow and Chen, 1992; Choe and Bulatao, 1992; WHO, 1994; Galway and Stover, 1995), yet, there has never been any attempt to incorporate the method-mix in measuring family planning performance.

The family planning performance assessment presented in this paper is based on the composite family planning performance index developed and used by Chaurasia (2020). This index takes into account the met demand for modern spacing methods; the met demand for permanent methods; and the method-mix. The index is a theoretical improvement over existing approaches of family planning performance measurement and has many advantages. It is easy to calculate and does not require any additional data. It can be calculated from method-specific prevalence rates and unmet need for spacing and limiting births that are readily available from existing population-based surveys such as Demographic and Health Surveys. The index ranges from 0 (poorest performance) to 1 (best performance) so as to permit spatial and temporal comparison of family planning performance. It also addresses the problem of perfect substitutability between the met demand for modern spacing methods and the met demand for permanent methods which is important in assessing family planning performance.

The paper is organized as follows. The family planning performance index proposed by Chaurasia (2020) is briefly described in the next section while section three describes the data source. The paper is based on survey-based estimates of the prevalence of different family planning methods and unmet need for spacing and limiting births made available by the United Nations Population Division. Section four discusses inter-country variation in contraceptive prevalence and unmet need for spacing and limiting. Section five analyses inter-country variation in family planning performance and the change in performance over time. The last section of the paper discusses the findings of the analysis in the context of the goals set under the FP2020 Initiative.

### Family Planning Performance Index

The family planning performance index, *p*, proposed by Chaurasia (2020) combines the met demand for modern spacing methods, met demand for permanent methods and method-mix into a composite index. The met demand for modern spacing methods is measured in terms of the index, *c*_*s*_, defined as

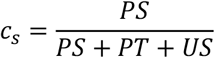

where *PS* is the prevalence of modern spacing methods; *PT* is the prevalence of traditional methods; and *US* is the unmet need for modern spacing methods. Here, it is assumed that users of traditional methods actually have an unmet need for modern spacing methods.

On the other hand, the met demand for permanent methods is measured in terms of the index, *c*_*p*_, which is defined as

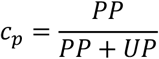

where *PP* is the prevalence of permanent methods; and *UP* is the unmet need for permanent methods of family planning.

It may be noticed that the sum of the met demand for modern spacing methods and the met demand for permanent methods is not equal to the met demand for family planning that has been identified under the FP2020 Initiative to monitor family planning progress. The met demand for family planning, *m*, by definition is given by

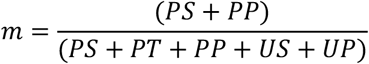

and it is easy to check that

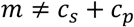

The indexes *c*_*s*_ and *c*_*p*_ may, be combined into the composite index *c* that reflects the demand for family planning in the following manner

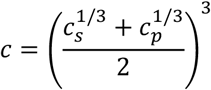

The index *c* is the power mean of the indexes *c*_*s*_ and *c*_*p*_. It places greater weight on that component (spacing or limiting) in which the met demand is low. For example, if *c*_*s*_ < *c*_*p*_, then the index *c* places greater weight on *c*_*s*_. On the other hand, if *c*_*s*_ > *c*_*p*_, it places greater weight on *c*_*p*_. When *c*_*s*_ = *c*_*p*_, the index assigns equal weight to *c*_*s*_ and *c*_*p*_. In this way, the index *c* penalizes those countries where there is an imbalance between the met demand of modern spacing methods and the met demand for permanent methods. It is obvious that the index *c* varies between 0 and 1 as both *c*_*s*_ and *c*_*p*_ vary between 0 and 1. It is also obvious that the higher the index *c*, the higher the met demand for family planning. The rationale of measuring the met demand for family planning in terms of the index *c* is that it provides impetus to improve performance in that component of family planning (spacing or limiting) in which the met demand is comparatively low.

On the other hand, an index reflecting the method-mix may be constructed on the basis of the method skew index proposed by Chaurasia (2020). This index is based on the concept of the dominance of one family planning method over others. The method skew index, *s*, is defined as

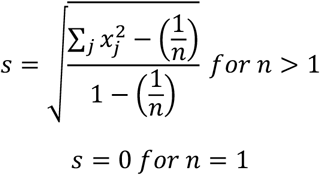

where *x*_*j*_ is the proportionate prevalence rate of the method *j* or the ratio of the prevalence of method *j* to all modern methods prevalence rate. The index *s* is independent of the number of family planning methods available and ranges between 0 and 1. When *s*=0, proportionate share of different methods in total family planning use is the same which implies that there is no method skew. When *s*=1, entire family planning use is confined to only one method so that the method-mix is completely skewed. Based on the index *s*, an index of method mix, *q*, may be defined as

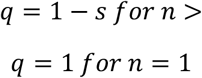

The lower the value of *q*, the higher the skewness in the method-mix and vice versa. When the proportionate share of different methods is the same in the total family planning use, the index *q* is zero and the higher the index *q* the more even or ‘balanced’ the proportionate distribution of total family planning use by different modern methods.

The family planning performance index, *p*, is now the composite of indexes *c* and *q*, and is defined as

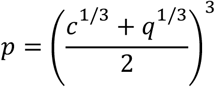

Since both *c* and *q* vary between 0 and 1, *p* also varies between 0 and 1. The higher the index *p* the higher the family planning performance in meeting the diverse family planning needs of women. From the performance perspective, it may be argued that indexes *c* and *q* should not be correlated while both *c* and *q* should be correlated with the performance index *p*. The data available from 113 countries suggests that inter-country variation in the index *c* (index *q*) explains only around 1 per cent of the inter-country variation in the index *q* (index *c*). On the other hand, inter-country variation in index *c* and in index *q* explains more than 98 per cent of per cent of the inter-country variation in the index *p*.

The index *p*, along with indexes *c* and *q*, constitutes a comprehensive family planning performance assessment framework that takes into account both the met demand for family planning and the composition of the met demand. Family planning performance may be rated as very poor if *p*<0.200; poor if 0.200≤*p*<0.400; average if 0.400≤*p*<0.600; good if 0.600≤*p*<0.800; and very good if *p*≥0.800. Family planning performance may also be characterized in terms of met demand for family planning and in terms of the method-mix.

### Data

The analysis is based on the database on world contraceptive use maintained by the United Nations Population Division (United Nations, 2020). This database is the compilation of 1202 nationally representative surveys that have been carried out in 196 countries or areas of the world sometimes during the period 1950 through 2019. The database maintained by the United Nations is regularly updated and the last update was done on 31 January 2020. The present analysis is, however, limited to only 113 countries that are selected on the basis of the following criteria: 1) the latest survey should have been carried out sometimes during the period 2010-2019; 2) estimates of the prevalence rate are available for different modern family planning methods separately; and 3) estimates of unmet need for family planning are available separately for spacing births and for limiting births married or in-union women aged 15-49 years. Out of the 113 countries covered in the present analysis, 47 countries are from Africa; 30 are from Asia; 20 are from Latin America and Caribbean; 11 are from Europe; and 5 are from the Pacific region of the world. The countries included in the present analysis also include 65 of the 69 lowest-income countries of the world that have been identified as focus countries under the FP2020 Initiative which has aimed at achieving the target of reaching an additional 120 million users of modern family planning methods in these countries by the year 2020 (FP2020, 2018).

### Prevalence of Modern Family Planning Methods

The latest estimates of the prevalence of modern family planning methods in 113 countries varies from a minimum of just 1.7 per cent in South Sudan (2010) to 77.4 per cent in Nicaragua (2011-12) with a median of 37.4 per cent and an inter-quartile range of 32.4 per cent (Table 1). In 29 countries, the prevalence of modern family planning methods is less than 20 per cent whereas, there is no country where the prevalence of modern methods of family planning is more than 80 per cent. There are only 15 countries where the prevalence of modern family planning methods ranges between 60-80 per cent. In 80 or more than 70 per cent countries, more than half of the married or in-union women aged 15-49 years have not been found using any modern family planning method according to the latest available survey estimates.

**Table 1.** Indicators of family planning use, unmet need for family planning and met demand for family planning in 114 countries

| Performance | Prevalence of modern family planning methods |  | Unmet need for family planning |  |  |  |  |  | Met demand for family planning |  |
| --- | --- | --- | --- | --- | --- | --- | --- | --- | --- | --- |
|  |  |  | All modern methods |  | For spacing births |  | For limiting births |  |  |  |
|  | Number of countries |  |  |  |  |  |  |  |  |  |
| Very poor | <0.2 | 29 | ≥0.20 | 50 | ≥0.20 | 14 | ≥0.20 | 0 | <0.2 | 4 |
| Poor | 0.2-0.4 | 34 | 0.15-0.2 | 22 | 0.15-0.2 | 13 | 0.15-0.2 | 6 | 0.2-0.4 | 27 |
| Average | 0.4-0.6 | 35 | 0.1-0.15 | 27 | 0.1-0.15 | 16 | 0.1-0.15 | 26 | 0.4-0.6 | 31 |
| Good | 0.6-0.8 | 15 | 0.05-0.1 | 13 | 0.05-0.1 | 38 | 0.05-0.1 | 56 | 0.6-0.8 | 37 |
| Very good | ≥0.8 | 0 | <0.05 | 1 | <0.05 | 22 | <0.05 | 25 | ≥0.8 | 14 |
|  | Summary measures of inter-country distribution |  |  |  |  |  |  |  |  |  |
| Minimum | 0.017 |  | 0.049 |  | 0.022 |  | 0.018 |  | 0.056 |  |
| Q1 | 0.198 |  | 0.125 |  | 0.054 |  | 0.057 |  | 0.394 |  |
| Median | 0.374 |  | 0.180 |  | 0.091 |  | 0.076 |  | 0.561 |  |
| Q3 | 0.520 |  | 0.259 |  | 0.164 |  | 0.106 |  | 0.725 |  |
| Maximum | 0.774 |  | 0.402 |  | 0.313 |  | 0.197 |  | 0.898 |  |
| IQR | 0.322 |  | 0.134 |  | 0.110 |  | 0.049 |  | 0.332 |  |
| CV | 0.513 |  | 0.431 |  | 0.597 |  | 0.457 |  | 0.369 |  |
| N | 113 |  | 113 |  | 113 |  | 113 |  | 113 |  |

Source: Author's calculations.

The unmet need for family planning (either spacing or limiting) also varies widely across 113 countries. There is only one country – Ukraine (2012) - where the unmet need for family planning is estimated to be less than 5 per cent. There are only 15 countries including Ukraine where the unmet need for family planning is estimated to be less than 10 per cent. By contrast, in Libya (2014), the unmet need for family planning is estimated to be more than 40 per cent, the highest among the 113 countries. There are 50 countries where the unmet need for family planning for either spacing or limiting births has been estimated to be at least 20 per cent.

The inter-country variation in the unmet need for spacing births, however, is different from that for limiting births. The unmet need for spacing births is found to be less than 5 per cent in 22 countries with the lowest unmet need for spacing births estimated in Peru (2018) followed by Viet Nam (2013-2014). On the other hand, the unmet need for spacing births is estimated to be more than 30 per cent in Libya (2014), the highest among 113 countries. There are 14 countries where the unmet need for spacing births is estimated to be more than 20 per cent. On the other hand, the unmet need for limiting births is less than 2 per cent in Ukraine (2012) but almost 20 per cent in Haiti (2012). There are only 6 countries where the unmet need for limiting births is more than 15 per cent whereas, there is no country where the unmet need for limiting births is 20 per cent or more. The inter-country coefficient of variation in the unmet need for spacing births is 0.597 as compared to inter-country coefficient of variation of 0.457 in the unmet need for limiting births. The inter-country variation in the unmet need for spacing births explains just around 6 per cent of the inter-country variation in the unmet need for limiting births and vice versa. This observation justifies considering, separately, unmet need for spacing births and unmet need for limiting births in measuring the met demand for family planning.

Combining the prevalence of modern family planning methods and the unmet need for family planning (either for spacing births or for limiting births), the met demand for family planning is estimated to vary from less than 6 per cent in South Sudan (2010) to almost 90 per cent in Thailand (2015-16), Democratic Republic of Korea (2017) and Nicaragua (2011-12).

There is, however, no country where the met demand for family planning is at least 90 per cent whereas there are only 14 countries where the met demand for family planning ranges between 80-90 per cent. In almost 40 per cent of the countries included in this analysis, the met demand for family planning remains less than 50 per cent which indicates that there is substantial scope of expanding the delivery of family planning services so as to meet the family planning demand of women which is very diverse.

### Family Planning Performance

#### Met Demand for Permanent Methods

The met demand for permanent methods is either very poor or poor (*c*_*p*_*<*0.400) in 79 or almost 70 per cent countries included in the present analysis. In seven countries – Benin (2017), Burkina Faso (2018), Côte d’Ivoire (2018), Ethiopia (2018), Guinea-Bissau (2018-19), Libya (2014), Sudan (2014) – the entire demand for permanent family planning methods remains unmet. There are only 10 countries where the met demand for permanent methods may be termed as very good (*c*_*p*_*≥*0.800) and 6 countries, where it may be termed as good (0.600≤*c*_*p*_*<*0.800). By contrast, the met demand for permanent methods is very poor (*c*_*p*_<0.200) in 54 countries.

#### Met Demand for Modern Spacing Methods

There are only 21 countries where the met demand for modern spacing method is either very poor or poor (*c*_*s*_*<*0.400) whereas, in 22 countries, it is very good (*c*_*s*_*≥*0.800). Unlike the met demand for permanent methods, there is no country where the met demand for modern spacing methods is zero and there are only 3 countries where it is very poor (*c*_*s*_*<*0.200). In majority of the countries, the met demand for modern spacing methods is higher than that for permanent methods. There are, however, notable exceptions such as India where the med demand for permanent methods is more than 83 per cent but the met demand for modern spacing methods is just around 50 per cent.

#### Met Demand Performance Index

Combining the met demand for permanent methods and the met demand for modern spacing methods, the met demand performance index *c*, reflecting family planning performance in terms of meeting the demand for family planning services varies from just around 0.034 in South Sudan (2010) to more than 0.900 in Nicaragua (2011). The family planning performance in terms of meeting the family planning demand of married or in-union women aged 15-49 years may be rated as very poor (*c*<0.200) in 32 countries and poor (0.200≤*c*<0.400) in 25 countries so that in more than half of the countries, family planning performance in terms of meeting the demand for family planning services remains far from satisfactory, either very poor or poor. There are only 9 countries – Nicaragua (2011-12), Cuba (2014), Dominican Republic (2014), El Salvador (2014), Thailand (2015-16), Colombia (2015-16), Mexico (2015), Costa Rica (2018), Bhutan (2010) - where family planning services delivery system appears to be able to meet at least 80 per of the demand for family planning as reflected through the index *c*. All but two of these 9 countries are in Latin America. In addition, there are 15 countries where family planning services delivery system appears to be able to meet 60-80 per cent of the demand for family planning. This means that there are only 24 countries where family planning performance in meeting family planning demand may be termed as satisfactory.

**Figure 1:**
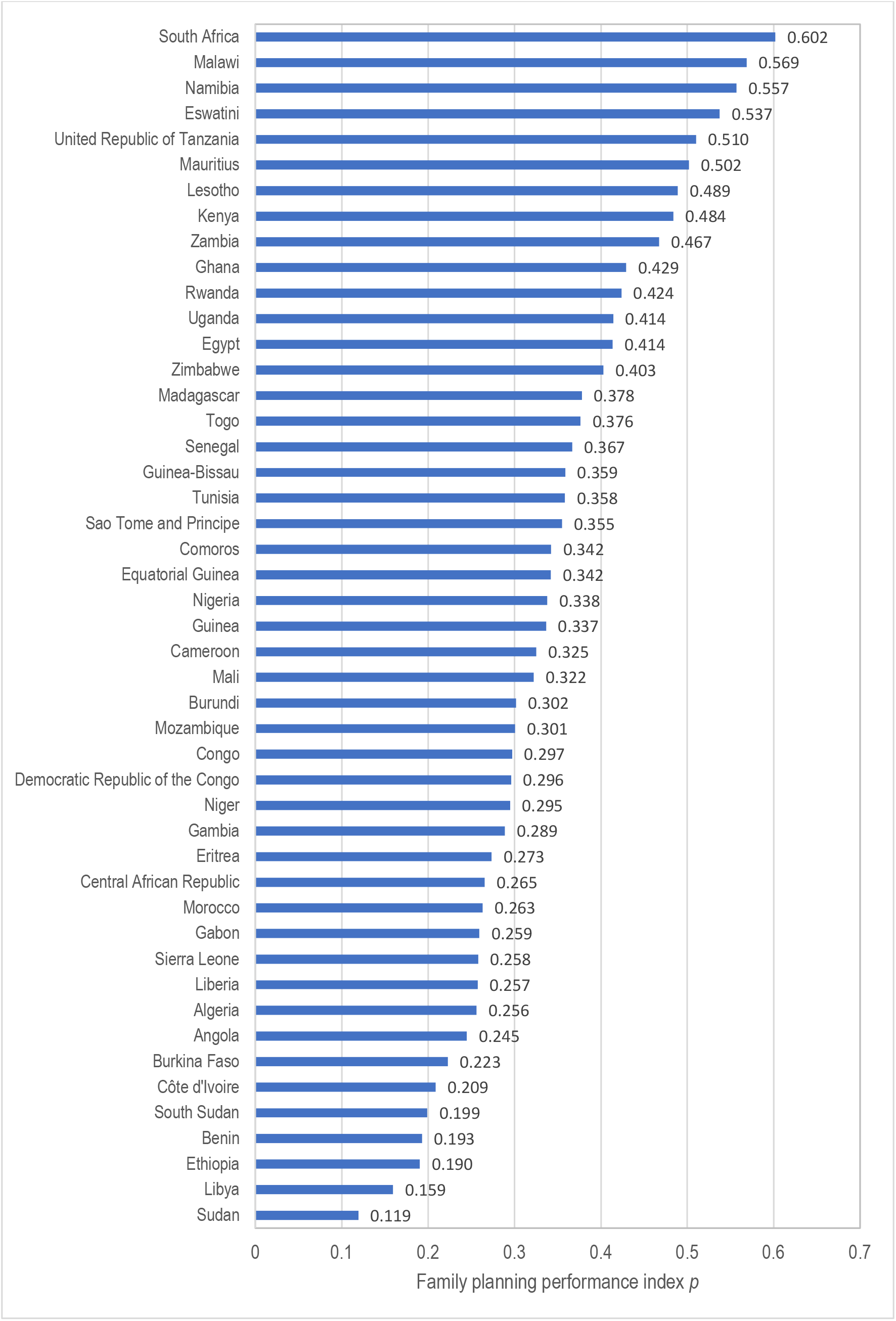
Family planning performance in 47 African Countries.

**Figure 2:**
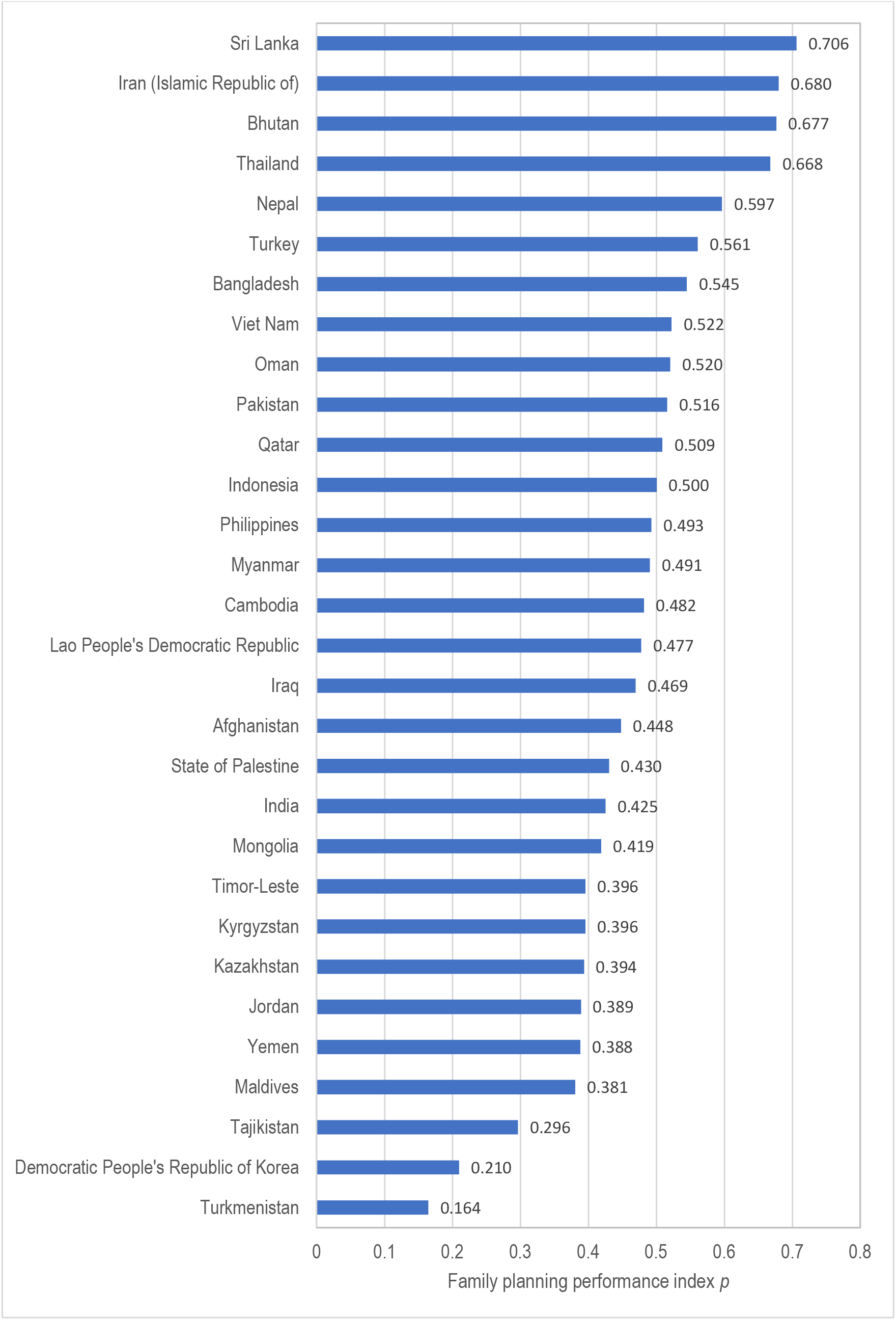
Family planning performance in 30 Asian countries.

**Figure 3:**
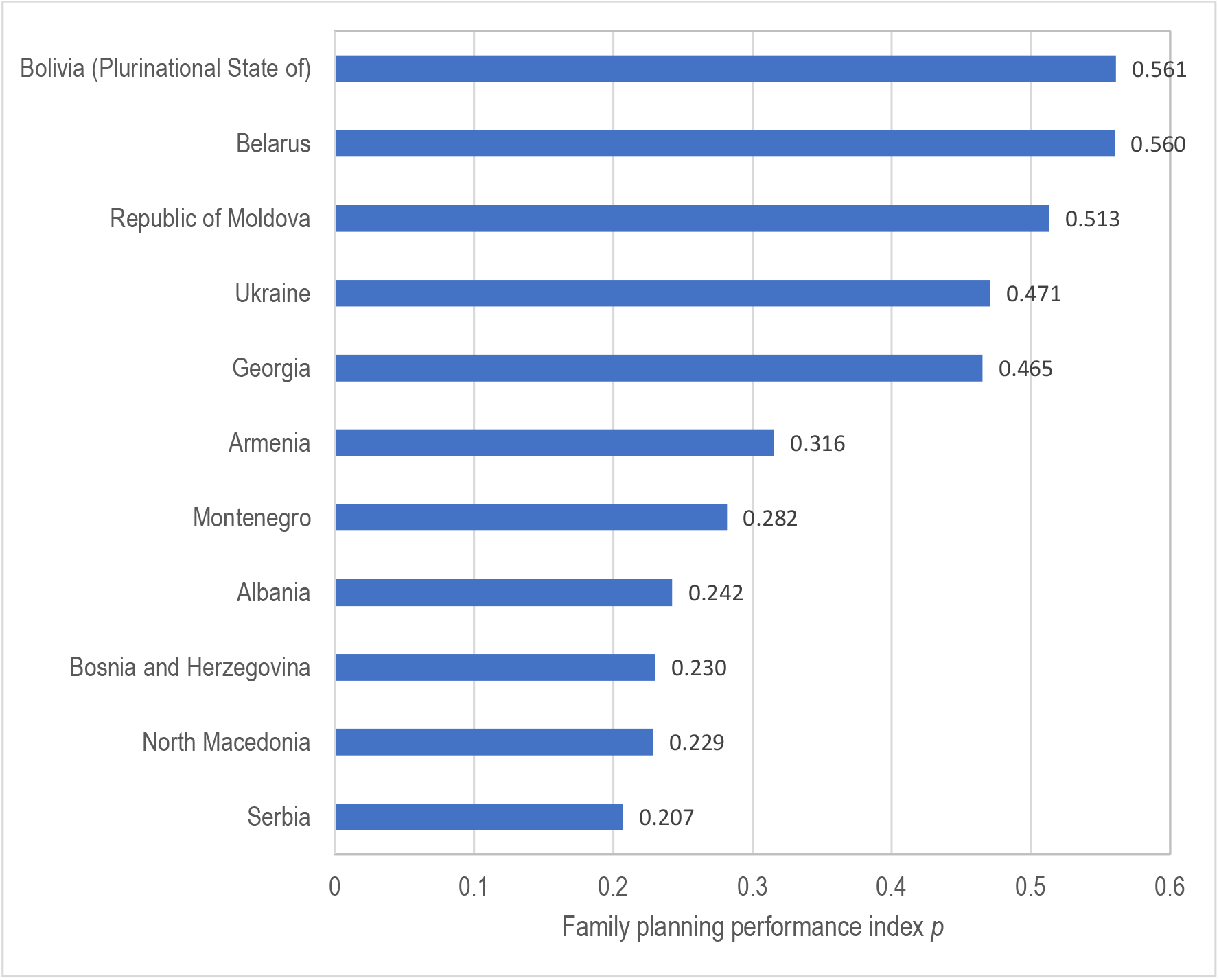
Family planning performance in 11 European countries.

**Figure 4:**
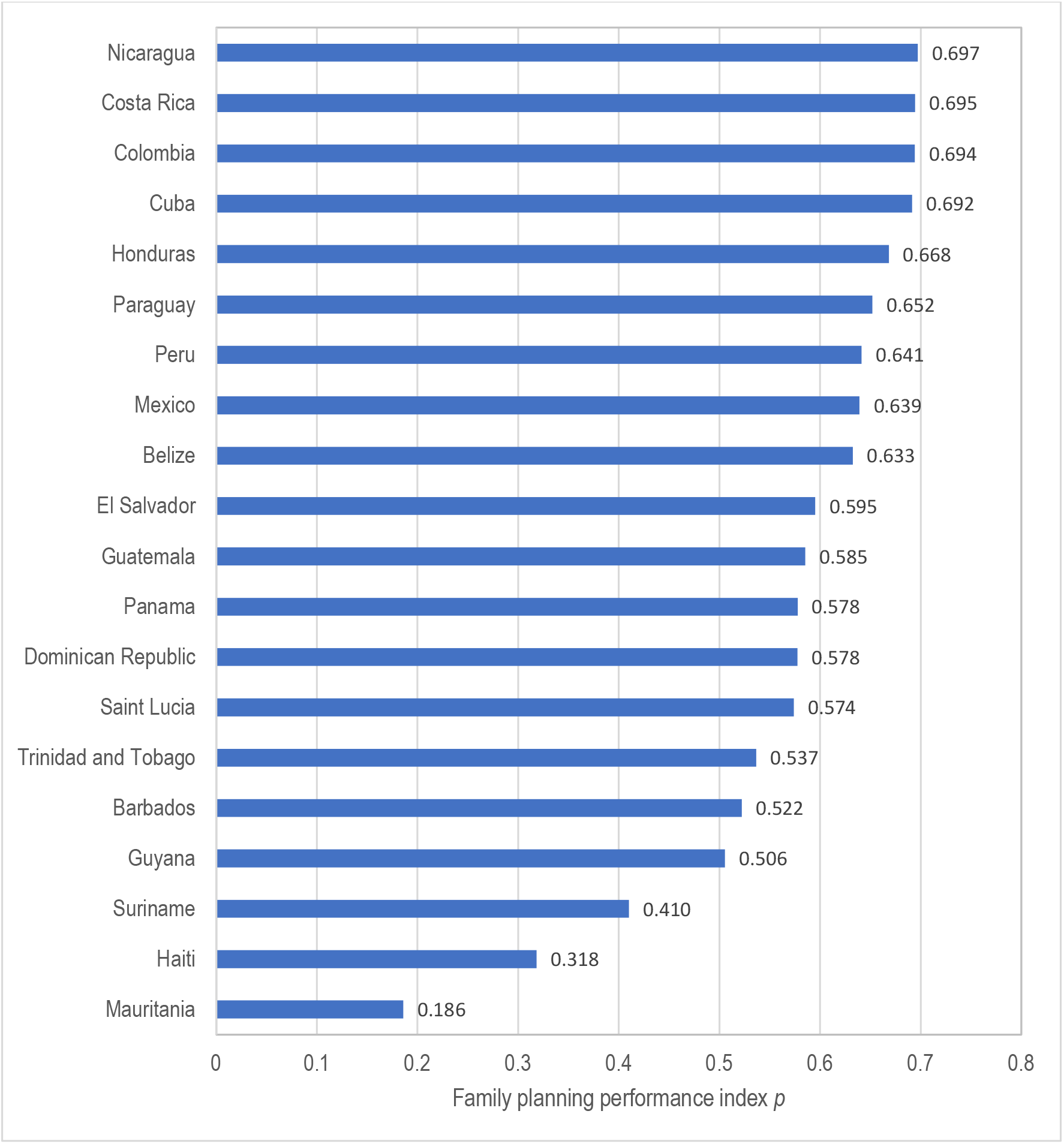
Family planning performance in 20 Latin American and Caribbean countries.

**Figure 5:**
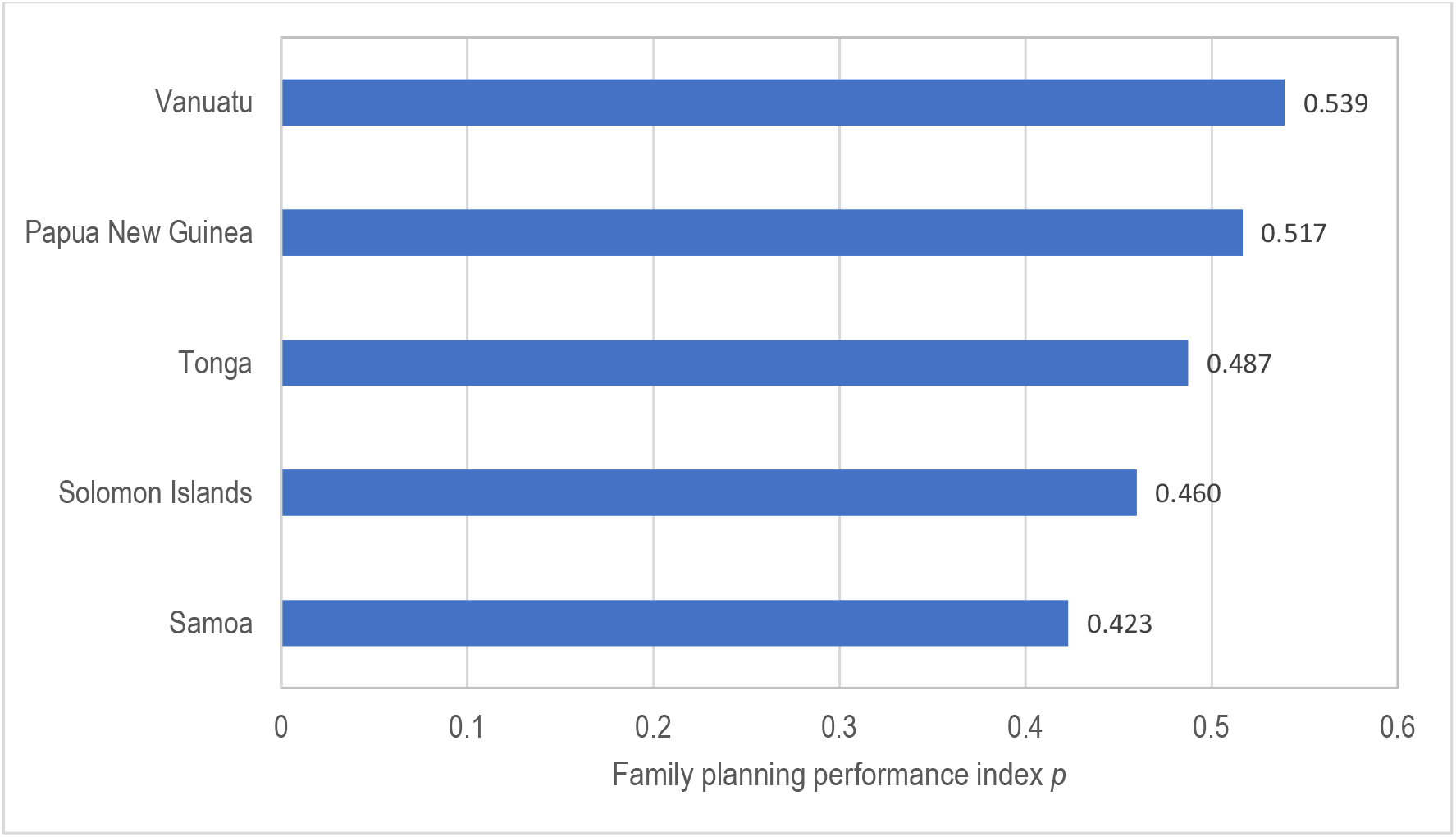
Family planning performance in 5 Oceanic countries.

#### Method-Mix

The method-mix index *q*, is found to be the lowest in Turkmenistan (2015) (*q*=0.071) but the lightest in Guinea-Bissau (2014) (*q*=0.689). In Turkmenistan, IUD, alone, accounts for more than 93 per cent of the total family planning use. Other countries where the method-mix index is very low are Morocco where 82 per cent of total family planning users are Pill users; Sudan where Pill accounts for almost 77 per cent of the total family planning use; and India where female sterilization accounts for more than 75 per cent of the total family planning use. There are 19 countries where family planning performance in terms of the method-mix may be termed as either poor or very poor (*q*<0.400) in these countries. In these countries, method-mix is heavily skewed as the total family planning use is heavily concentrated in one method only. The method-mix remains skewed in all the countries included in the present analysis as there is no country where the index *q* is at least 0.800. There are only 13 countries where the method-mix may be termed as comparatively balanced as the index *q* ranges between 0.600-0.800 in these countries.

#### Family Planning Performance

Taking into account both family planning performance in terms of meeting family planning needs of women as reflected through the index *c* and the family planning performance in terms of method-mix as reflected through the index *p*, the family planning performance, reflected through the index, *p*, ranges from 0.119 in Sudan (2014) to 0.706 in Sri Lanka (2016) with a median of 0.424 and an IQR or 0.236. There is no country where family planning performance can be termed as very good (*p*≥0.800) whereas there are only 14 countries where the family planning performance may be termed as good (0.600≤*p*<0.800).

These countries are: South Africa (2016), Belize (2011), Mexico (2015), Peru (2018), Paraguay (2016), Thailand (2015-16), Honduras (2011-12), Bhutan (2010), Iran (2010-11), Cuba (2014), Colombia (2015-16), Costa Rica (2018), Nicaragua (2011-12) and Sri Lanka (2016). Sri Lanka is the only country where the family planning performance index is estimated to be more than 0.700. On the other hand, family planning performance is poor in 43 countries (0.200≤*p*<0.400) and very poor in 7 countries (*p*<0.200) – Sudan (2014), Libya (2014), Turkmenistan (2015-16), Mauritania (2015), Ethiopia (2018), Benin (2018) and South Sudan (2010).

It may also be observed from table 2 that when no distinction is made between the met demand for spacing and the met demand for limiting births, there are 51 countries where family planning performance may be termed as good or very good in terms of meeting the demand for family planning. However, when a distinction is made between the met demand for modern spacing methods and the met demand for permanent methods, then there are only 24 countries where the family planning performance may be termed as good or very good. It appears that a high met demand for family planning in a number of countries is due to the perfect substitutability between the met demand for modern spacing methods and the met demand for permanent methods which is implicit when the met demand is estimated without making a distinction between the met demand for modern spacing methods and the met demand for permanent methods.

**Table 2.** Family planning performance in 114 countries, 2010-2019.

| Performance | Met demand for |  | Performance index |  |  |
| --- | --- | --- | --- | --- | --- |
| | Permanent methods | Modern spacing methods | $c$ | $q$ | $p$ |
| Number of countries |  |  |  |  |  |
| Very poor (<0.200) | 54 | 3 | 32 | 4 | 7 |
| Poor (0.200-0.400) | 25 | 18 | 25 | 15 | 43 |
| Average (0.400-0.600) | 18 | 33 | 32 | 81 | 49 |
| Good (0.600-0.800) | 6 | 37 | 15 | 13 | 14 |
| Very good ($\geq 0.800$) | 10 | 22 | 9 | 0 | 0 |
| Summary measures of inter-country distribution |  |  |  |  |  |
| Minimum | 0.000 | 0.049 | 0.034 | 0.054 | 0.119 |
| Q1 | 0.066 | 0.450 | 0.185 | 0.429 | 0.301 |
| Median | 0.209 | 0.611 | 0.398 | 0.500 | 0.424 |
| Q3 | 0.465 | 0.767 | 0.549 | 0.552 | 0.534 |
| Maximum | 0.921 | 0.939 | 0.902 | 0.689 | 0.706 |
| IQR | 0.399 | 0.317 | 0.364 | 0.122 | 0.236 |
| CV | 0.919 | 0.343 | 0.576 | 0.234 | 0.345 |
| N | 113 | 113 | 113 | 113 | 113 |

Source: Author's calculations.

**Table 3.** Family planning performance improvement in 87 countries

| Performance improvement | Average annual per cent change (*AAPC*) in |  |  |  |  |
| --- | --- | --- | --- | --- | --- |
| | $c_p$ | $c_s$ | $c$ | $q$ | $p$ |
|  | Number of countries |  |  |  |  |
| Performance decreased ($AAPC < 0$) | 34 | 19 | 31 | 38 | 30 |
| Marginal improvement ($0 \leq AAPC < 1.0$) | 10 | 17 | 12 | 21 | 21 |
| Mild improvement ($1.0 \leq AAPC < 2.0$) | 4 | 17 | 12 | 12 | 22 |
| Moderate improvement ($2.0 \leq AAPC < 3.0$) | 6 | 8 | 10 | 4 | 5 |
| Substantial improvement ($AAPC \geq 3.0$) | 33 | 26 | 22 | 12 | 9 |
|  | Summary measures of inter-country distribution |  |  |  |  |
| Minimum | -36.82 | -6.20 | -19.73 | -4.34 | -9.14 |
| Q1 | -2.62 | 0.09 | -0.91 | -0.75 | -0.43 |
| Median | 0.93 | 1.38 | 1.00 | 0.15 | 0.53 |
| Q3 | 5.26 | 3.34 | 2.99 | 1.36 | 1.42 |
| Maximum | 98.29 | 16.20 | 19.87 | 12.80 | 10.25 |
| IQR | 7.88 | 3.25 | 3.90 | 2.12 | 1.85 |
| CV | 4.20 | 1.70 | 4.01 | 4.50 | 4.62 |
| N | 87 | 87 | 87 | 87 | 87 |

Source: Author's calculations

Table 2 also suggests that, in most of the countries, the method-mix remains skewed, dominated by either one or two methods only which also reflects a negative feature of family planning performance. A skewed method-mix implies that family planning services are essentially directed towards satisfying family planning needs of a particular section of women only and family planning needs of other women remain largely unmet. A focus on only one or two methods of family planning may also be the reason for a low met demand for family planning. If the family planning services are to meet the family planning needs of all married or in-union women aged 15-49 years, then, it is imperative, that the method-mix should not be dominated by specific family planning methods or it should not be skewed but more balanced.

### Trend in Family Planning Performance

The change in the family planning performance index, *p*, during a temporal segment (*t*_*2*_>*t*_*1*_), can be measured in terms of annual per cent change (*APC*) under the assumption that the change is constant during the temporal segment. The *APC* is calculated as

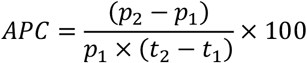

The *APC* in different temporal segments of a given time period provides a complete characterization of the trend in family planning performance over the given time period. It may, however, be noticed that the *APC* in different temporal segments of a time period may not be the same and the relative contribution of *APC* in a given temporal segment to the change in the entire period is a function of the length of the temporal segment. A high *APC* in a short temporal segment may have only a small contribution to the change over the entire time period. The *APCs* in different temporal segments of a given time period can, however, be combined to obtain average annual per cent change (*AAPC*) during the entire time period. *AAPC* is the weighted average of different *APCs* within the time period with weights equal to the proportionate length of different temporal segments. *AAPC* serves as a single summary measure of the change during the entire time period in which *APCs* in different temporal segments are essentially different and the length of different temporal segments is also different.

Information about method specific prevalence and unmet need for spacing and limiting births for at least two points in time during the period 2000-2019 is available for 87 of the 113 countries included in the analysis. In 30 countries, the *AAPC* in the index *p* has been negative indicating that family planning performance in these countries has decreased over time. The decrease in the index *p* has been the most rapid in Panama during 2013-15 when the index *p* decreased by more than 9 per cent per year. The decrease in family planning performance has also been quite rapid in Sierra Leone (2013-2016), Serbia (2010-2014), Azerbaijan (2001-2006), Tunisia (2011-2018) and Suriname (2010-2018) indicating a decrease in family planning performance as the index *p* decreased by more than 3 per cent per year. There are, in fact, only 9 countries – Montenegro (2013-2018), Malawi (2000-2016), Rwanda (2000-2015), Congo (2005-2015), Togo (2010-2014), Democratic Republic of Congo (2007-2014), Oman (2007-2014), Ukraine (2007-2012) and Timor-Leste (2009-2016) – where improvement in family planning performance can be termed as substantial as *AAAC* in the index *p* has been 3 per cent per year and more in these countries with the highest *AAPC* observed in Timor-Leste (2009-2016). In rest of the countries, the *AAAC* in the index *p* has, at best, been marginal which suggests that there has been only a marginal improvement in family planning performance in these countries.

The change in the performance index *p*, is the result of the change in indexes *c* and *q*. The index *c* has decreased in 31 countries with the most rapid decrease observed in Panama during 2013-2015. At the same time, there are 22 countries where the index *c* increased by at least 3 per cent per year with the most rapid increase observed in Oman during 2007-2014. There are, however, only three countries in addition to Oman where the increase in the index *c* has been at least 10 per cent per year. These countries are: Togo during 2010-2014; Rwanda during 2000-2015; and Ukraine during 2007-2012. In majority of the countries, however, the increase in the index *c* has been marginal which indicating only a marginal improvement in family planning performance in terms of meeting the family planning needs of women.

The method-mix index *q*, on the other hand, decreased in 38 countries which indicates that the skewness in the method-mix in these countries has increased over time. There are only 12 countries where the method-mix index *q* increased at a rate of at least 3 per cent per year with Timor-Leste the only country where the index *q* increased at a rate of more than 10 per cent per year during 2009-2016 indicating a rapid decrease in the skewness in the mix. The proportion of total family planning users using injectable in Timor-Leste decreased from more than 76 per cent during 2009-10 to around 49 per cent during 2016 whereas users of implant increased from just around 4 per cent to more than 26 per cent during this period so that the index *q* increased from 0.253 to almost 0.480 during 2009-16. In majority of the countries, however, there has been only a marginal improvement in the index of method-mix over time indicating only a marginal improvement in family planning performance in the context of a balanced method-mix to meet the diverse family planning needs of women.

The change in the index *c* that reflects the extent to which the demand for family planning services is met, is the result of the change in the met demand for permanent methods and the change in the met demand for modern spacing methods of family planning. The family performance has been different in terms of the met demand for modern spacing methods and the met demand for permanent methods. There are only 45 countries where the met demand for both permanent methods and modern spacing methods increased over time though at varying rates of improvement. On the other hand, there are 11 countries, where met demand for both permanent methods and modern spacing methods decreased over time. In 23 countries, the met demand for permanent methods decreased but the met demand for modern spacing methods decreased over time. This leaves only 8 countries where the met demand for modern spacing methods decreased but the met demand for permanent methods increased. This means that, in 34 countries the met demand for permanent methods decreased whereas the met demand for modern spacing methods decreased in only 19 countries. At the same time, the met demand for permanent methods increased at a rate of at least 3 per cent per year in 33 countries. but in 26 countries in case of modern spacing methods. In most of the countries, family planning performance in terms of the improvement in the met demand for permanent methods has been different from the performance in terms of the improvement in the met demand for modern spacing methods reflecting the imbalance in the performance in terms of meeting the diverse family planning needs of women.

## Discussions and Conclusions

In this paper, we have presented a two-dimensional perspective of family planning performance in 113 countries considering both the extent up to which the family planning needs of married or in-union women aged 15-49 are met the balance in the method-mix. The analysis is probably and so obviously the first that considers both the demand for family planning and the composition of family planning use by different family planning methods in assessing the family planning performance. The analysis reveals that, in general, family planning performance in most of the countries remains far from satisfactory in terms of satisfying the family planning needs of married or in-union women aged 15-49 years. There is also evidence to suggest that, in majority of the countries, the method-mix has turned more skewed over time indicating the increased dependence of the family planning services delivery system on only one method to meet the diverse family planning needs of women which is not a welcome feature of family planning performance. Similarly, family planning performance in terms of meeting the demand for modern spacing methods and in terms of meeting the demand for permanent methods has also been different in most of the countries suggesting that the improvement in family planning performance has not been balanced in most of the countries. These observations suggest that there are only a small number of countries where family planning performance may be termed as satisfactory in meeting the diverse family planning needs of women. In most of the countries, there is substantial scope for improvement in family planning performance in terms of either meeting the demand for modern spacing methods or meeting the demand for permanent methods or in terms of achieving a balanced method mix that has been advocated from the perspective of rights-based family planning. What is even more concerned is the observation that, in a substantial proportion of countries, family planning performance has worsened in at least one of the three dimensions of family planning services delivery.

The family planning movement in the world is now almost seven decades old. The launch of the first official family planning program by India way back in 1952 may be taken as the beginning of this movement. The genesis of the movement was grounded in the proposition that fertility regulation and curtailing population growth through family planning would contribute significantly towards addressing a range of development concerns facing the developing countries. Following this premise, substantial efforts have been made, resources mobilized and commitments made to main-stream family planning in the development discourse of most of the developing countries. Family planning has now universally been accepted as an integral component of any strategy directed towards meeting reproductive health needs of the people, especially women and is valued as a reproductive right. It has also been accepted as an important development intervention that has implications for broader development goals such as United Nations Sustainable Development Goals. The FP2020 Initiative, launched in 2012, has set an ambitious target of recruiting 120 million new acceptors of family planning by 2020 (120 by 2020) in the context of meeting the pressing reproductive health care needs of women, especially, in 69 focus countries. This target, however, could not be achieved (FP2020, 2020) because of the unsatisfactory family planning performance in most of the countries, as revealed through the present analysis. It appears that despite specific commitments at the global level, most of the countries have not been able to mobilize additional resources necessary to ensure universal access to family planning services necessary to meet the reproductive health needs of women and in a large number of countries, family planning performance, instead of improving, has deteriorated over time. In most of the countries, family planning is largely a prerogative of official family planning efforts. This means that a reinvigoration of official family planning efforts is urgently needed so as to effectively meet the reproductive health needs of women.

## Data Availability

Data used in this study have been taken from database on world contraceptive use maintained by United Nations Population Division. All data are available online.

https://www.un.org/en/development/desa/population/publications/dataset/contraception/wcu2019.asp#:~:text=The%20data%20set%2C%20World%20Contraceptive,available%20as%20of%20February%202019.&text=Both%20datasets%20are%20used%20for,1.

---

**Appendix Table 1.** Family planning use in 114 countries, 2010-2019

| Country | Year | CPR | CPRm | UM | Met demand |
| --- | --- | --- | --- | --- | --- |
| Africa |  |  |  |  |  |
| Algeria | 2012-2013 | 0.571 | 0.495 | 0.070 | 0.772 |
| Angola | 2015-2016 | 0.136 | 0.125 | 0.380 | 0.242 |
| Benin | 2017-2018 | 0.156 | 0.120 | 0.323 | 0.251 |
| Burkina Faso | 2018-2019 | 0.325 | 0.307 | 0.233 | 0.550 |
| Burundi | 2016-2017 | 0.285 | 0.224 | 0.297 | 0.385 |
| Cameroon | 2014 | 0.343 | 0.210 | 0.180 | 0.402 |
| Central African Republic | 2010-2011 | 0.152 | 0.121 | 0.270 | 0.287 |
| Comoros | 2012 | 0.194 | 0.142 | 0.316 | 0.278 |
| Congo | 2014-2015 | 0.301 | 0.185 | 0.179 | 0.385 |
| Côte d'Ivoire | 2018 | 0.233 | 0.196 | 0.265 | 0.394 |
| Democratic Republic of the Congo | 2013-2014 | 0.204 | 0.075 | 0.277 | 0.156 |
| Egypt | 2014 | 0.585 | 0.569 | 0.126 | 0.800 |
| Equatorial Guinea | 2011 | 0.126 | 0.095 | 0.338 | 0.205 |
| Eritrea | 2010 | 0.083 | 0.070 | 0.274 | 0.196 |
| Eswatini | 2014 | 0.661 | 0.655 | 0.152 | 0.806 |
| Ethiopia | 2018 | 0.401 | 0.378 | 0.206 | 0.623 |
| Gabon | 2012 | 0.311 | 0.194 | 0.265 | 0.337 |
| Gambia | 2018 | 0.167 | 0.163 | 0.265 | 0.377 |
| Ghana | 2017 | 0.330 | 0.274 | 0.263 | 0.462 |
| Guinea | 2018 | 0.110 | 0.103 | 0.177 | 0.359 |
| Guinea-Bissau | 2014 | 0.160 | 0.144 | 0.223 | 0.376 |
| Kenya | 2017 | 0.605 | 0.587 | 0.149 | 0.778 |
| Lesotho | 2018 | 0.650 | 0.646 | 0.160 | 0.798 |
| Liberia | 2013 | 0.202 | 0.191 | 0.311 | 0.372 |
| Libya | 2014 | 0.277 | 0.163 | 0.402 | 0.240 |
| Madagascar | 2017 | 0.478 | 0.386 | 0.164 | 0.601 |
| Malawi | 2015-2016 | 0.592 | 0.581 | 0.187 | 0.746 |
| Mali | 2018 | 0.172 | 0.164 | 0.239 | 0.399 |
| Mauritius | 2014 | 0.638 | 0.320 | 0.125 | 0.419 |
| Morocco | 2018 | 0.708 | 0.591 | 0.113 | 0.720 |
| Mozambique | 2015 | 0.271 | 0.253 | 0.231 | 0.504 |
| Namibia | 2013 | 0.561 | 0.553 | 0.175 | 0.751 |
| Niger | 2017 | 0.189 | 0.181 | 0.222 | 0.441 |
| Nigeria | 2018 | 0.167 | 0.121 | 0.189 | 0.340 |
| Rwanda | 2014-2015 | 0.533 | 0.475 | 0.189 | 0.658 |
| Sao Tome and Principe | 2014 | 0.405 | 0.374 | 0.337 | 0.504 |
| Senegal | 2017 | 0.278 | 0.262 | 0.219 | 0.527 |
| Sierra Leone | 2016 | 0.225 | 0.218 | 0.263 | 0.447 |
| South Africa | 2016 | 0.546 | 0.540 | 0.149 | 0.777 |
| South Sudan | 2010 | 0.040 | 0.017 | 0.263 | 0.056 |
| Sudan | 2014 | 0.122 | 0.117 | 0.266 | 0.302 |
| Togo | 2017 | 0.239 | 0.215 | 0.340 | 0.371 |
| Tunisia | 2018 | 0.504 | 0.443 | 0.199 | 0.630 |
| Uganda | 2017 | 0.385 | 0.339 | 0.294 | 0.500 |
| United Republic of Tanzania | 2015-2016 | 0.384 | 0.320 | 0.221 | 0.529 |
| Zambia | 2013-2014 | 0.491 | 0.447 | 0.211 | 0.637 |
| Zimbabwe | 2015 | 0.668 | 0.658 | 0.104 | 0.852 |
| Asia |  |  |  |  |  |
| Afghanistan | 2015-2016 | 0.225 | 0.198 | 0.245 | 0.421 |
| Bangladesh | 2014 | 0.623 | 0.539 | 0.120 | 0.725 |
| Bhutan | 2010 | 0.656 | 0.654 | 0.117 | 0.846 |
| Cambodia | 2014 | 0.563 | 0.388 | 0.125 | 0.564 |
| Democratic People's Republic of Korea | 2017 | 0.702 | 0.688 | 0.066 | 0.896 |
| India | 2015-2016 | 0.535 | 0.478 | 0.129 | 0.720 |
| Indonesia | 2016-2017 | 0.610 | 0.588 | 0.148 | 0.775 |
| Iran (Islamic Republic of) | 2010-2011 | 0.787 | 0.570 | 0.057 | 0.675 |
| Iraq | 2018 | 0.528 | 0.361 | 0.143 | 0.538 |
| Jordan | 2017-2018 | 0.518 | 0.374 | 0.142 | 0.567 |
| Kazakhstan | 2018 | 0.530 | 0.501 | 0.155 | 0.732 |
| Kyrgyzstan | 2018 | 0.394 | 0.378 | 0.190 | 0.647 |
| Lao People's Democratic Republic | 2017 | 0.540 | 0.490 | 0.143 | 0.717 |
| Maldives | 2016-2017 | 0.186 | 0.147 | 0.314 | 0.294 |
| Mongolia | 2018 | 0.481 | 0.452 | 0.228 | 0.638 |
| Myanmar | 2015-2016 | 0.523 | 0.513 | 0.162 | 0.749 |
| Nepal | 2016-2017 | 0.526 | 0.428 | 0.237 | 0.561 |
| Oman | 2014 | 0.297 | 0.188 | 0.178 | 0.396 |
| Pakistan | 2017-2018 | 0.342 | 0.250 | 0.173 | 0.485 |
| Philippines | 2017 | 0.541 | 0.401 | 0.167 | 0.566 |
| Qatar | 2012 | 0.375 | 0.344 | 0.124 | 0.689 |
| Sri Lanka | 2016 | 0.617 | 0.513 | 0.075 | 0.741 |
| State of Palestine | 2014 | 0.572 | 0.441 | 0.109 | 0.648 |
| Tajikistan | 2017 | 0.293 | 0.271 | 0.227 | 0.521 |
| Thailand | 2015-2016 | 0.783 | 0.755 | 0.062 | 0.893 |
| Timor-Leste | 2016 | 0.261 | 0.237 | 0.253 | 0.461 |
| Turkey | 2018 | 0.700 | 0.490 | 0.116 | 0.600 |
| Turkmenistan | 2015-2016 | 0.502 | 0.471 | 0.121 | 0.756 |
| Viet Nam | 2013-2014 | 0.758 | 0.570 | 0.061 | 0.696 |
| Yemen | 2013 | 0.335 | 0.252 | 0.287 | 0.405 |
| Europe |  |  |  |  |  |
| Albania | 2017-2018 | 0.461 | 0.037 | 0.151 | 0.060 |
| Armenia | 2015-2016 | 0.572 | 0.280 | 0.125 | 0.402 |
| Belarus | 2012 | 0.631 | 0.520 | 0.070 | 0.742 |
| Bolivia (Plurinational State of) | 2016 | 0.664 | 0.451 | 0.232 | 0.503 |
| Bosnia and Herzegovina | 2011-2012 | 0.458 | 0.120 | 0.090 | 0.219 |
| Georgia | 2018 | 0.406 | 0.327 | 0.231 | 0.513 |
| Montenegro | 2018 | 0.207 | 0.116 | 0.210 | 0.278 |
| North Macedonia | 2011 | 0.402 | 0.128 | 0.172 | 0.223 |
| Republic of Moldova | 2012 | 0.594 | 0.417 | 0.095 | 0.605 |
| Serbia | 2014 | 0.584 | 0.184 | 0.149 | 0.251 |
| Ukraine | 2012 | 0.655 | 0.478 | 0.049 | 0.679 |
| Latin America and Caribbean |  |  |  |  |  |
| Barbados | 2012 | 0.592 | 0.554 | 0.199 | 0.700 |
| Belize | 2011 | 0.552 | 0.520 | 0.159 | 0.731 |
| Colombia | 2015-2016 | 0.809 | 0.759 | 0.067 | 0.866 |
| Costa Rica | 2018 | 0.709 | 0.690 | 0.137 | 0.816 |
| Cuba | 2014 | 0.738 | 0.722 | 0.080 | 0.883 |
| Dominican Republic | 2014 | 0.695 | 0.680 | 0.114 | 0.841 |
| El Salvador | 2014 | 0.719 | 0.676 | 0.111 | 0.814 |
| Guatemala | 2014-2015 | 0.606 | 0.489 | 0.139 | 0.656 |
| Guyana | 2014 | 0.338 | 0.325 | 0.280 | 0.526 |
| Haiti | 2012 | 0.345 | 0.313 | 0.353 | 0.448 |
| Honduras | 2011-2012 | 0.732 | 0.638 | 0.107 | 0.760 |
| Mauritania | 2015 | 0.178 | 0.156 | 0.336 | 0.304 |
| Mexico | 2015 | 0.665 | 0.646 | 0.130 | 0.813 |
| Nicaragua | 2011-2012 | 0.804 | 0.774 | 0.058 | 0.898 |
| Panama | 2014-2015 | 0.508 | 0.469 | 0.242 | 0.625 |
| Paraguay | 2016 | 0.684 | 0.665 | 0.121 | 0.826 |
| Peru | 2018 | 0.763 | 0.550 | 0.063 | 0.666 |
| Saint Lucia | 2011-2012 | 0.555 | 0.525 | 0.170 | 0.724 |
| Suriname | 2018 | 0.391 | 0.385 | 0.284 | 0.570 |
| Trinidad and Tobago | 2011 | 0.403 | 0.376 | 0.243 | 0.582 |
| Oceania |  |  |  |  |  |
| Papua New Guinea | 2016-2018 | 0.367 | 0.305 | 0.259 | 0.487 |
| Samoa | 2014 | 0.270 | 0.243 | 0.348 | 0.393 |
| Solomon Islands | 2015 | 0.293 | 0.243 | 0.347 | 0.380 |
| Tonga | 2012 | 0.341 | 0.284 | 0.252 | 0.479 |
| Vanuatu | 2013 | 0.489 | 0.371 | 0.242 | 0.508 |

Source: Author's calculations based on United Nations (2020)
CPR Contraceptive prevalence rate
CPRm Modern methods prevalence rate
UM Unmet need for family planning

**Appendix Table 2.** Family planning performance in 114 countries, 2010-2019

| Country | Period | $c_p$ | $c_s$ | $c$ | $q$ | $p$ |
| --- | --- | --- | --- | --- | --- | --- |
| Africa |  |  |  |  |  |  |
| Algeria | 2012-2013 | 0.185 | 0.798 | 0.420 | 0.141 | 0.256 |
| Angola | 2015-2016 | 0.008 | 0.315 | 0.086 | 0.531 | 0.245 |
| Benin | 2017-2018 | 0.000 | 0.313 | 0.040 | 0.541 | 0.193 |
| Burkina Faso | 2018-2019 | 0.000 | 0.595 | 0.075 | 0.494 | 0.223 |
| Burundi | 2016-2017 | 0.042 | 0.494 | 0.185 | 0.460 | 0.302 |
| Cameroon | 2014 | 0.027 | 0.464 | 0.154 | 0.590 | 0.325 |
| Central African Republic | 2010-2011 | 0.029 | 0.337 | 0.126 | 0.481 | 0.265 |
| Comoros | 2012 | 0.090 | 0.316 | 0.180 | 0.581 | 0.342 |
| Congo | 2014-2015 | 0.041 | 0.424 | 0.164 | 0.488 | 0.297 |
| Côte d'Ivoire | 2018 | 0.000 | 0.434 | 0.055 | 0.524 | 0.209 |
| Democratic Republic of the Congo | 2013-2014 | 0.104 | 0.164 | 0.132 | 0.560 | 0.296 |
| Egypt | 2014 | 0.129 | 0.901 | 0.398 | 0.429 | 0.414 |
| Equatorial Guinea | 2011 | 0.077 | 0.254 | 0.148 | 0.657 | 0.342 |
| Eritrea | 2010 | 0.029 | 0.237 | 0.099 | 0.584 | 0.273 |
| Eswatini | 2014 | 0.282 | 0.906 | 0.535 | 0.540 | 0.537 |
| Ethiopia | 2018 | 0.000 | 0.709 | 0.089 | 0.348 | 0.190 |
| Gabon | 2012 | 0.072 | 0.383 | 0.187 | 0.349 | 0.259 |
| Gambia | 2018 | 0.046 | 0.434 | 0.174 | 0.446 | 0.289 |
| Ghana | 2017 | 0.123 | 0.547 | 0.284 | 0.617 | 0.429 |
| Guinea | 2018 | 0.039 | 0.428 | 0.163 | 0.602 | 0.337 |
| Guinea-Bissau | 2014 | 0.029 | 0.454 | 0.156 | 0.689 | 0.359 |
| Kenya | 2017 | 0.269 | 0.850 | 0.505 | 0.463 | 0.484 |
| Lesotho | 2018 | 0.181 | 0.901 | 0.449 | 0.531 | 0.489 |
| Liberia | 2013 | 0.032 | 0.450 | 0.159 | 0.389 | 0.257 |
| Libya | 2014 | 0.000 | 0.276 | 0.035 | 0.434 | 0.159 |
| Madagascar | 2017 | 0.182 | 0.668 | 0.374 | 0.382 | 0.378 |
| Malawi | 2015-2016 | 0.582 | 0.798 | 0.684 | 0.467 | 0.569 |
| Mali | 2018 | 0.053 | 0.475 | 0.193 | 0.498 | 0.322 |
| Mauritius | 2014 | 0.465 | 0.399 | 0.431 | 0.580 | 0.502 |
| Morocco | 2018 | 0.120 | 0.788 | 0.356 | 0.188 | 0.263 |
| Mozambique | 2015 | 0.029 | 0.580 | 0.186 | 0.455 | 0.301 |
| Namibia | 2013 | 0.444 | 0.831 | 0.617 | 0.500 | 0.557 |
| Niger | 2017 | 0.042 | 0.467 | 0.177 | 0.456 | 0.295 |
| Nigeria | 2018 | 0.029 | 0.416 | 0.146 | 0.652 | 0.338 |
| Rwanda | 2014-2015 | 0.144 | 0.733 | 0.362 | 0.491 | 0.424 |
| Sao Tome and Principe | 2014 | 0.037 | 0.662 | 0.219 | 0.537 | 0.355 |
| Senegal | 2017 | 0.068 | 0.589 | 0.242 | 0.529 | 0.367 |
| Sierra Leone | 2016 | 0.011 | 0.545 | 0.141 | 0.426 | 0.258 |
| South Africa | 2016 | 0.485 | 0.872 | 0.660 | 0.547 | 0.602 |
| South Sudan | 2010 | 0.014 | 0.070 | 0.034 | 0.598 | 0.199 |
| Sudan | 2014 | 0.000 | 0.370 | 0.047 | 0.243 | 0.119 |
| Togo | 2017 | 0.083 | 0.457 | 0.219 | 0.594 | 0.376 |
| Tunisia | 2018 | 0.089 | 0.786 | 0.321 | 0.398 | 0.358 |
| Uganda | 2017 | 0.202 | 0.572 | 0.356 | 0.479 | 0.414 |
| United Republic of Tanzania | 2015-2016 | 0.347 | 0.566 | 0.447 | 0.578 | 0.510 |
| Zambia | 2013-2014 | 0.209 | 0.700 | 0.406 | 0.534 | 0.467 |
| Zimbabwe | 2015 | 0.154 | 0.903 | 0.424 | 0.383 | 0.403 |
| Asia |  |  |  |  |  |  |
| Afghanistan | 2015-2016 | 0.205 | 0.470 | 0.319 | 0.607 | 0.448 |
| Bangladesh | 2014 | 0.468 | 0.778 | 0.610 | 0.485 | 0.545 |
| Bhutan | 2010 | 0.741 | 0.903 | 0.819 | 0.552 | 0.677 |
| Cambodia | 2014 | 0.307 | 0.609 | 0.441 | 0.525 | 0.482 |
| Democratic People's Republic of Korea | 2017 | 0.265 | 0.939 | 0.533 | 0.054 | 0.210 |
| India | 2015-2016 | 0.834 | 0.502 | 0.654 | 0.257 | 0.425 |
| Indonesia | 2016-2017 | 0.311 | 0.864 | 0.541 | 0.461 | 0.500 |
| Iran (Islamic Republic of) | 2010-2011 | 0.890 | 0.616 | 0.745 | 0.619 | 0.680 |
| Iraq | 2018 | 0.270 | 0.594 | 0.411 | 0.533 | 0.469 |
| Jordan | 2017-2018 | 0.161 | 0.632 | 0.345 | 0.437 | 0.389 |
| Kazakhstan | 2018 | 0.117 | 0.855 | 0.372 | 0.416 | 0.394 |
| Kyrgyzstan | 2018 | 0.173 | 0.724 | 0.385 | 0.407 | 0.396 |
| Lao People's Democratic Republic | 2017 | 0.331 | 0.811 | 0.536 | 0.424 | 0.477 |
| Maldives | 2016-2017 | 0.242 | 0.325 | 0.281 | 0.501 | 0.381 |
| Mongolia | 2018 | 0.191 | 0.762 | 0.413 | 0.426 | 0.419 |
| Myanmar | 2015-2016 | 0.309 | 0.888 | 0.549 | 0.437 | 0.491 |
| Nepal | 2016-2017 | 0.564 | 0.557 | 0.561 | 0.634 | 0.597 |
| Oman | 2014 | 0.444 | 0.371 | 0.407 | 0.653 | 0.520 |
| Pakistan | 2017-2018 | 0.533 | 0.463 | 0.497 | 0.535 | 0.516 |
| Philippines | 2017 | 0.404 | 0.623 | 0.506 | 0.480 | 0.493 |
| Qatar | 2012 | 0.271 | 0.758 | 0.474 | 0.546 | 0.509 |
| Sri Lanka | 2016 | 0.758 | 0.735 | 0.746 | 0.667 | 0.706 |
| State of Palestine | 2014 | 0.281 | 0.686 | 0.454 | 0.408 | 0.430 |
| Tajikistan | 2017 | 0.066 | 0.659 | 0.258 | 0.338 | 0.296 |
| Thailand | 2015-2016 | 0.895 | 0.893 | 0.894 | 0.483 | 0.668 |
| Timor-Leste | 2016 | 0.189 | 0.507 | 0.322 | 0.480 | 0.396 |
| Turkey | 2018 | 0.578 | 0.607 | 0.592 | 0.531 | 0.561 |
| Turkmenistan | 2015-2016 | 0.078 | 0.817 | 0.316 | 0.071 | 0.164 |
| Viet Nam | 2013-2014 | 0.446 | 0.717 | 0.571 | 0.477 | 0.522 |
| Yemen | 2013 | 0.148 | 0.495 | 0.287 | 0.510 | 0.388 |
| Europe |  |  |  |  |  |  |
| Albania | 2017-2018 | 0.103 | 0.049 | 0.073 | 0.571 | 0.242 |
| Armenia | 2015-2016 | 0.099 | 0.435 | 0.227 | 0.425 | 0.316 |
| Belarus | 2012 | 0.500 | 0.766 | 0.624 | 0.501 | 0.560 |
| Bolivia (Plurinational State of) | 2016 | 0.370 | 0.561 | 0.459 | 0.677 | 0.561 |
| Bosnia and Herzegovina | 2011-2012 | 0.034 | 0.241 | 0.106 | 0.426 | 0.230 |
| Georgia | 2018 | 0.221 | 0.638 | 0.393 | 0.545 | 0.465 |
| Montenegro | 2018 | 0.054 | 0.376 | 0.167 | 0.440 | 0.282 |
| North Macedonia | 2011 | 0.056 | 0.270 | 0.136 | 0.356 | 0.229 |
| Republic of Moldova | 2012 | 0.484 | 0.624 | 0.551 | 0.477 | 0.513 |
| Serbia | 2014 | 0.036 | 0.289 | 0.122 | 0.325 | 0.207 |
| Ukraine | 2012 | 0.333 | 0.692 | 0.491 | 0.451 | 0.471 |
| Latin America and Caribbean |  |  |  |  |  |  |
| Barbados | 2012 | 0.316 | 0.792 | 0.518 | 0.526 | 0.522 |
| Belize | 2011 | 0.735 | 0.729 | 0.732 | 0.543 | 0.633 |
| Colombia | 2015-2016 | 0.919 | 0.820 | 0.868 | 0.545 | 0.694 |
| Costa Rica | 2018 | 0.751 | 0.864 | 0.806 | 0.594 | 0.695 |
| Cuba | 2014 | 0.845 | 0.903 | 0.874 | 0.537 | 0.692 |
| Dominican Republic | 2014 | 0.907 | 0.757 | 0.830 | 0.383 | 0.578 |
| El Salvador | 2014 | 0.894 | 0.735 | 0.812 | 0.421 | 0.595 |
| Guatemala | 2014-2015 | 0.797 | 0.576 | 0.681 | 0.500 | 0.585 |
| Guyana | 2014 | 0.218 | 0.630 | 0.388 | 0.645 | 0.506 |
| Haiti | 2012 | 0.075 | 0.611 | 0.256 | 0.390 | 0.318 |
| Honduras | 2011-2012 | 0.834 | 0.725 | 0.778 | 0.569 | 0.668 |
| Mauritania | 2015 | 0.010 | 0.374 | 0.103 | 0.304 | 0.186 |
| Mexico | 2015 | 0.814 | 0.812 | 0.813 | 0.492 | 0.639 |
| Nicaragua | 2011-2012 | 0.921 | 0.884 | 0.902 | 0.526 | 0.697 |
| Panama | 2014-2015 | 0.474 | 0.767 | 0.609 | 0.548 | 0.578 |
| Paraguay | 2016 | 0.563 | 0.890 | 0.714 | 0.594 | 0.652 |
| Peru | 2018 | 0.709 | 0.657 | 0.683 | 0.602 | 0.641 |
| Saint Lucia | 2011-2012 | 0.447 | 0.802 | 0.607 | 0.542 | 0.574 |
| Suriname | 2018 | 0.272 | 0.655 | 0.436 | 0.385 | 0.410 |
| Trinidad and Tobago | 2011 | 0.372 | 0.687 | 0.513 | 0.560 | 0.537 |
| Oceania |  |  |  |  |  |  |
| Papua New Guinea | 2016-2018 | 0.395 | 0.538 | 0.463 | 0.574 | 0.517 |
| Samoa | 2014 | 0.280 | 0.473 | 0.368 | 0.483 | 0.423 |
| Solomon Islands | 2015 | 0.395 | 0.370 | 0.383 | 0.546 | 0.460 |
| Tonga | 2012 | 0.537 | 0.434 | 0.484 | 0.491 | 0.487 |
| Vanuatu | 2013 | 0.477 | 0.522 | 0.499 | 0.581 | 0.539 |

Source: Author's calculations

**Appendix Table 3.** Trend in family planning performance

| Country | Period |  | Average annual per cent change (*AAPC*)<br>in |  |  |  |  |
| --- | --- | --- | --- | --- | --- | --- | --- |
| | | | $c_p$ | $c_s$ | $c$ | $q$ | $p$ |
| Africa |  |  |  |  |  |  |  |
| Algeria | 2006 | 2013 | 3.82 | -0.31 | 1.12 | -1.92 | -0.22 |
| Benin | 2001 | 2018 | -5.38 | 4.04 | -1.85 | -0.96 | -1.73 |
| Burkina Faso | 2003 | 2019 | 42.72 | 8.37 | 5.84 | -1.15 | -0.14 |
| Burundi | 2010 | 2017 | -4.46 | 4.44 | 0.99 | 1.61 | 1.34 |
| Cameroon | 2004 | 2014 | -11.67 | 4.72 | -3.21 | 3.33 | 0.35 |
| Central African Republic | 1994 | 2011 | -3.67 | 14.17 | 2.96 | -0.97 | 0.23 |
| Congo | 2005 | 2015 | 2.51 | 7.93 | 5.71 | 3.78 | 4.54 |
| Côte d'Ivoire | 2011 | 2018 | 7.03 | 4.16 | -3.20 | 3.41 | 0.17 |
| Democratic Republic of the Congo | 2007 | 2014 | 0.28 | 6.53 | 3.14 | 6.35 | 5.07 |
| Egypt | 2000 | 2014 | 0.90 | 0.03 | 0.20 | 1.70 | 0.91 |
| Eritrea | 2002 | 2010 | 0.00 | -0.19 | -0.13 | 0.63 | 0.35 |
| Eswatini | 2006 | 2014 | 4.59 | 1.40 | 2.10 | -0.62 | 0.51 |
| Ethiopia | 2000 | 2018 | 17.15 | 8.82 | 4.68 | -0.36 | 0.37 |
| Gabon | 2000 | 2012 | -2.85 | 5.02 | 1.02 | -2.74 | -1.37 |
| Gambia | 2013 | 2018 | -11.54 | 12.90 | -0.18 | -2.01 | -1.26 |
| Ghana | 2003 | 2017 | 13.35 | 3.53 | 4.72 | 0.04 | 1.27 |
| Guinea | 2005 | 2018 | 9.82 | 7.18 | 7.86 | 0.16 | 2.12 |
| Kenya | 2003 | 2017 | 0.33 | 3.50 | 2.09 | -0.76 | 0.57 |
| Lesotho | 2004 | 2018 | 4.09 | 1.51 | 2.30 | 0.21 | 1.17 |
| Liberia | 2006 | 2013 | -5.17 | 8.98 | 2.85 | -2.92 | -0.84 |
| Madagascar | 2003 | 2017 | 7.29 | 3.06 | 4.54 | -1.09 | 1.29 |
| Malawi | 2000 | 2016 | 6.87 | 3.12 | 4.75 | 1.57 | 3.13 |
| Mali | 2001 | 2018 | 10.28 | 4.97 | 4.41 | -0.26 | 1.37 |
| Morocco | 2003 | 2018 | -3.66 | -0.14 | -1.80 | -2.26 | -2.01 |
| Mozambique | 2003 | 2015 | -3.66 | 1.02 | -1.09 | -0.83 | -1.25 |
| Namibia | 2000 | 2013 | 0.93 | 0.79 | 0.84 | -0.41 | 0.22 |
| Niger | 2006 | 2017 | -2.18 | 2.17 | -1.30 | 0.03 | -0.64 |
| Nigeria | 2003 | 2018 | 4.92 | 1.72 | 1.28 | 0.13 | 0.47 |
| Rwanda | 2000 | 2015 | 11.32 | 16.20 | 12.47 | -1.12 | 3.57 |
| Sao Tome and Principe | 2006 | 2014 | 4.34 | -0.39 | -0.12 | 5.77 | 3.00 |
| Senegal | 2005 | 2017 | 6.80 | 6.90 | 5.41 | -0.58 | 1.69 |
| Sierra Leone | 2013 | 2016 | -26.74 | 6.27 | -8.98 | -4.34 | -6.37 |
| South Africa | 2003 | 2016 | -1.69 | -0.28 | -0.97 | 0.91 | -0.14 |
| Togo | 2010 | 2017 | 36.08 | 4.02 | 10.83 | 1.32 | 4.75 |
| Tunisia | 2011 | 2018 | -11.66 | 0.53 | -6.78 | -0.75 | -4.24 |
| Uganda | 2000 | 2017 | 4.35 | 2.69 | 3.02 | -0.89 | 0.75 |
| United Republic of Tanzania | 2004 | 2016 | 3.67 | 2.54 | 3.03 | 0.87 | 1.85 |
| Zambia | 2001 | 2014 | 3.96 | 3.18 | 3.48 | 0.29 | 1.63 |
| Zimbabwe | 2005 | 2015 | -2.48 | 0.49 | -0.67 | 3.78 | 1.36 |
| Asia |  |  |  |  |  |  |  |
| Bangladesh | 2004 | 2014 | 2.08 | 1.17 | 1.16 | 1.30 | 1.19 |
| | | | $c_p$ | $c_s$ | $c$ | $q$ | $p$ |
| Cambodia | 2000 | 2014 | 11.21 | 3.54 | 5.95 | -0.87 | 1.94 |
| India | 2005 | 2016 | 0.20 | 1.85 | 0.90 | 0.62 | 0.73 |
| Indonesia | 2002 | 2017 | -0.12 | 0.01 | -0.09 | -0.45 | -0.28 |
| Iraq | 2011 | 2018 | -6.09 | 0.67 | -2.94 | 1.06 | -1.05 |
| Jordan | 2002 | 2018 | -2.71 | -0.07 | -1.30 | 0.10 | -0.60 |
| Kazakhstan | 2010 | 2018 | 4.07 | -0.52 | -0.34 | 3.10 | 1.14 |
| Kyrgyzstan | 2012 | 2018 | -2.78 | 1.08 | -0.86 | 3.57 | 1.11 |
| Lao People's Democratic Republic | 2000 | 2017 | 6.10 | 1.48 | 3.15 | -1.02 | 0.91 |
| Maldives | 2009 | 2017 | -6.40 | -3.19 | -4.90 | -0.63 | -2.82 |
| Mongolia | 2003 | 2018 | -0.01 | -0.30 | -2.00 | 0.01 | -1.29 |
| Myanmar | 2001 | 2016 | -0.30 | 1.70 | 0.79 | -0.83 | -0.05 |
| Nepal | 2001 | 2017 | 0.11 | 1.40 | 0.71 | 1.14 | 0.83 |
| Oman | 2007 | 2014 | 35.78 | 9.57 | 19.87 | 0.29 | 6.66 |
| Pakistan | 2000 | 2018 | 5.22 | 0.80 | 2.78 | -0.43 | 1.01 |
| Philippines | 2003 | 2017 | -0.34 | 2.02 | 0.81 | -0.68 | -0.02 |
| Sri Lanka | 2000 | 2016 | 1.06 | 2.70 | 1.82 | 1.18 | 1.48 |
| State of Palestine | 2010 | 2014 | -3.64 | -0.16 | -1.73 | -0.46 | -1.11 |
| Tajikistan | 2012 | 2017 | 5.36 | 0.48 | 1.88 | 2.75 | 2.33 |
| Timor-Leste | 2009 | 2016 | 24.23 | 1.26 | 7.71 | 12.80 | 10.25 |
| Turkey | 2003 | 2018 | 1.85 | 0.72 | 1.13 | 0.42 | 0.75 |
| Turkmenistan | 2015 | 2016 | -3.73 | 0.21 | -1.55 | -2.97 | -2.16 |
| Viet Nam | 2002 | 2014 | -1.87 | 0.59 | -0.71 | 3.37 | 1.09 |
| Yemen | 2006 | 2013 | -2.42 | 1.73 | -0.14 | 0.23 | 0.06 |
| Europe |  |  |  |  |  |  |  |
| Albania | 2008 | 2018 | -6.38 | -6.20 | -6.30 | 0.16 | -2.86 |
| Armenia | 2000 | 2016 | 13.46 | 2.37 | 1.50 | -1.08 | -0.14 |
| Bolivia (Plurinational State of) | 2003 | 2016 | 2.37 | 1.17 | 1.68 | 0.48 | 1.02 |
| Montenegro | 2013 | 2018 | 98.29 | -3.12 | 8.92 | -0.30 | 3.01 |
| Republic of Moldova | 2005 | 2012 | 4.67 | 0.78 | 2.49 | 1.46 | 1.98 |
| Serbia | 2010 | 2014 | -8.33 | -4.18 | -5.70 | -4.03 | -4.75 |
| Ukraine | 2007 | 2012 | 57.78 | 0.58 | 14.39 | 1.89 | 7.29 |
| Latin America and Caribbean |  |  |  |  |  |  |  |
| Colombia | 2000 | 2016 | 0.68 | 1.35 | 1.00 | -0.13 | 0.46 |
| Costa Rica | 2011 | 2018 | -2.22 | -0.50 | -1.38 | 0.49 | -0.53 |
| Cuba | 2010 | 2014 | 1.75 | -0.62 | 0.51 | 0.15 | 0.35 |
| Dominican Republic | 2002 | 2014 | 0.13 | 1.38 | 0.72 | 2.13 | 1.29 |
| Guatemala | 2002 | 2015 | 2.75 | 3.04 | 2.86 | 0.19 | 1.51 |
| Guyana | 2009 | 2014 | 0.07 | -3.07 | -1.86 | 1.11 | -0.33 |
| Haiti | 2000 | 2012 | -2.53 | 2.04 | 0.24 | -1.57 | -0.92 |
| Honduras | 2005 | 2012 | 2.92 | 1.28 | 2.10 | 0.19 | 1.16 |
| Mauritania | 2007 | 2015 | 20.14 | 2.94 | 2.72 | 1.41 | 1.92 |
| Mexico | 2009 | 2015 | -1.52 | 1.65 | 0.00 | 6.84 | 2.78 |
| Nicaragua | 2001 | 2012 | 2.29 | 0.60 | 1.42 | -0.75 | 0.39 |
| Panama | 2013 | 2015 | -36.82 | 0.04 | -19.73 | 3.90 | -9.14 |
| | | | $c_p$ | $c_s$ | $c$ | $q$ | $p$ |
| Peru | 2000 | 2018 | 1.20 | 0.37 | 0.77 | -0.26 | 0.25 |
| Suriname | 2010 | 2018 | -6.35 | -2.47 | -4.44 | -2.05 | -3.37 |
| Trinidad and Tobago | 2006 | 2011 | 5.30 | -1.30 | 1.34 | -0.23 | 0.53 |
| Oceania |  |  |  |  |  |  |  |
| Samoa | 2009 | 2014 | 8.29 | 0.13 | 3.40 | 2.11 | 2.71 |
| Solomon Islands | 2006 | 2015 | -5.37 | -2.79 | -4.25 | 2.07 | -1.59 |

Source: Author's calculations

